# A multistage mixed-methods evaluation protocol for the national testing response during the COVID-19 pandemic in England

**DOI:** 10.1101/2022.10.27.22281604

**Authors:** Reshania Naidoo, Ben Lambert, Merryn Voysey, Rima Shretta, Claire Keene, Marta Wanat, Billie Andersen-Waine, Prabin Dahal, Kasia Stepniewska, Rachel Hounsell, Sassy Molyneux, Emily Rowe, Sarah Pinto-Duschinsky, Gulsen Yenidogan, Tom Fowler, Lisa J White, the EY-Oxford Health Analytics Consortium

**Author notes:** Corresponding author: Reshania Naidoo. The EY-Oxford Health Analytics Consortium membership list is attached in Appendix A.

## Abstract

**Introduction:** In 2020, the UK government established a large-scale testing programme to rapidly identify individuals in England who were infected with SARS-CoV-2 and had COVID-19. This comprised part of the UK government’s COVID-19 response strategy, to protect those at risk of severe COVID-19 disease and death and to reduce the burden on the health system. To assess the success of this approach, the UK Health Security Agency (UKHSA) commissioned an independent evaluation of the activities delivered by the National Health System (NHS) testing programme in England. The primary purpose of this evaluation will be to capture key learnings from the rollout of testing to different target populations via various testing services between October 2020 and March 2022 and to use these insights to formulate recommendations for future pandemic preparedness strategy. In this protocol, we detail the rationale, approach and study design.

**Methods and analysis:** The proposed study involves a stepwise mixed-methods approach, aligned with established methods for the evaluation of complex interventions in health, to retrospectively assess the combined impact of key asymptomatic and symptomatic testing services nationally. The research team will first develop a Theory of Change, formulated in collaboration with testing service stakeholders, to understand the causal pathways and intended and unintended outcomes of each testing service and explore contextual impacts on each testing service’s intended outcomes. Insights gained will help identify indicators to evaluate how the combined aims of the testing programme were achieved, using a mixed methods approach.

**Ethics and dissemination:** The study protocol was granted ethics approval by the UKHSA Research Ethics and Governance Group (reference NR0347). All relevant ethics guidelines will be followed throughout. Findings arising from this evaluation will be used to inform lessons learnt and recommendations for UKHSA on appropriate pandemic preparedness testing programme designs; findings will also be disseminated in peer-reviewed journals and at academic conferences. This will be the first evaluation to produce a portfolio of evidence in relation to the testing effectiveness and public health impact of the national testing programme in England, encompassing behavioural, economic, equity and public health impacts. These findings will strengthen the evidence base with regards to the effectiveness of COVID-19 testing and identify which aspects are necessary to prioritise in mitigating future pandemic threats when deploying a complex public health intervention such as testing.

**Transparency declaration:** The lead author (the manuscript’s guarantor) affirms that the manuscript is an honest, accurate and transparent account of the study being reported; no important aspects of the study have been omitted, and any discrepancies from the study as planned have been explained.

**Strengths and limitations of this protocol:**

- Strengths of this mixed methods evaluation protocol include the use of theory-based, complex evaluation approaches and an iterative and participatory approach with the stakeholder (UKHSA) to the evaluation process.
- Given the scale and complexity of the COVID-19 testing response in England, there is a scarcity of previous relevant research, either in England or appropriate international comparators, warranting the mixed methods evaluation approach we will employ.
- To the best of the authors’ knowledge, this is the first national-scale evaluation of the COVID-19 testing programme in England to incorporate the broadest scope of testing services, a programme that formed an integral part of the UK pandemic response strategy. The approach proposed could be applied to the evaluation of pandemic responses in other contexts or to other types of interventions.
- Whereas most complex interventions are ideally accompanied by a prospective evaluation design initiated at the time of the intervention or earlier, this study will predominantly comprise a retrospective evaluation and is therefore limited by the quality of existing research and the data available to the research team at the time of conducting the evaluation, within the specified eight-month period allocated by UKHSA. As the UK government is in the process of consolidating data and policy related to the COVID-19 pandemic and subject to an independent inquiry, certain datasets may not be available to the researchers at the time of conducting the evaluation.
- The scope of testing services to be evaluated and the selection of methods has been guided by the study sponsor team within UKHSA and must be achievable within the timeframe of the funding allocated to the study (eight months). Therefore, some trade-offs had to be made in terms of selecting research methods that would be feasible within this time constraint. For future evaluations, a mixed methods approach could be complemented by qualitative interviews with members of the public to gauge their experiences of testing and test-related behaviours, as well as an evaluation of other testing services that were out of scope for this research, including in prisons, the private sector and the events testing programme.

## Introduction

On 11 March 2020, the World Health Organization (WHO) declared COVID-19 a pandemic and exhorted member states to ‘test, test, test’.(1) In response to the COVID-19 pandemic, the UK government committed to mass testing, with initial testing commencing in March 2020.(2) NHS Test and Trace (NHSTT) was formally established in in May 2020, as a part of the Department of Health and Social Care (DHSC), to lead an ‘at scale’ national testing and tracing service.(3) The UK Health Security Agency (UKHSA) was established, also as an Executive Agency of DHSC, on 1 April 2021 and was operational on 1 October 2021.(4) UKHSA combines the health protection, clinical and scientific functions formerly carried out by Public Health England (PHE) with the functions of NHSTT and the Joint Biosecurity Centre (JBC).(5)

During the initial phases of the pandemic, in the absence of pharmaceutical interventions or a vaccine, testing was seen as a key means to reopen society. The UK government’s COVID-19 response strategy was therefore deployed in the context of mitigating the impact of the pandemic and the measures employed to control it on key areas of society, such as education, protection of livelihoods and preventative and mental health.(6) NHSTT was tasked with providing mass-scale testing and tracing systems to rapidly identify individuals with COVID-19 and their close contacts, thereby minimising the spread of the disease.(7) The NHSTT programme had four main stated objectives: 1) to increase the speed and availability of testing, 2) to identify close contacts of positive cases and require them to isolate, 3) to contain local outbreaks via a coordinated response, and 4) to enable the government to learn more about the virus and explore ways to ease infection control measures as the science developed.(3) The programme, at its scale, was the first of its kind in the UK and was created and delivered at pace during a period of unprecedented uncertainty.

The testing programme component of NHSTT played an integral role in the government’s COVID-19 response through the programme’s various testing services. The testing programme sought to work in partnership with national and local public health bodies, local authorities, the NHS, and commercial and academic providers. The delivery of testing for each of the target populations was multi-modal, through combinations of in-person testing, e.g. public regional testing sites and mobile testing units, and via pharmacies and home direct self-test kit deliveries; it was also driven in part by the technology available at the time, e.g. accredited self-sample collection was originally not an option, so physical testing sites were required. The delivery of testing was initially focused on regional and local PCR testing sites, followed by service-specific testing sites, then PCR and LFD home testing was rolled out as evidence accrued that this was a viable approach. These approaches to testing were subject to ongoing revision by policymakers throughout the pandemic, dependent on factors such as changing epidemiological prevalence, the emergence of new variants of concern, e.g. the Delta and Omicron variants, updated scientific evidence and vaccination rollout.(8)

Tests for COVID-19 include those that detect the presence of the SARS-CoV-2 virus and those that detect the presence of antibodies to the virus.(9) Tests for the virus that detect viral nucleic acid, such as polymerase chain reaction (PCR) and loop-mediated isothermal amplification (LAMP), are usually performed in a laboratory. These tests can appear positive beyond the period of infectiousness.(10) Lateral flow devices (LFDs) are test devices that can detect SARS-CoV-2 viral protein (antigen) and represent a more rapid approach to testing. They can also be used for self-testing, although they are less sensitive than nucleic acid-based tests (11) and tend only to appear positive during the period of maximal viral shedding.(12) At the start of the pandemic, only PCR tests were available for testing swabs from suspected cases of COVID-19; however, LFDs for COVID-19 were rapidly developed and being evaluated for use by mid-2020.(13) LFDs were initially rolled out for asymptomatic testing, followed by a confirmatory PCR test in the case of a positive LFD result.

The overall success and effectiveness of any national COVID-19 testing service is dependent on multiple contextual factors and the combined impacts of the various testing services. From a public health perspective, increased testing has been shown to result in reduced transmission of SARS-CoV-2 and associated hospitalisations and mortality.(14-18) Behavioural responses to testing strategies are dependent on public awareness and trust, with adherence to testing policy shown to be driven by perceptions of disease risk and socioeconomic factors.(19) From an equity perspective, COVID-19 testing has been found to exacerbate existing health inequities, with disparities in access to testing for individuals from ethnic minority backgrounds and those living in socially deprived areas.(20) The economic impact of testing can be assessed via cost-effectiveness evaluations of testing strategies, including testing unit costs, operational deployment costs and, on a macro-economic level, quantifying the economic productivity gained from shortened isolation periods and savings to the taxpayer.(21, 22)

UKHSA has appointed a consortium from Ernst & Young LLP in partnership with academics from the University of Oxford, contracted through Oxford University Innovation Limited, to carry out an evaluation of the testing programme; the evaluation will take eight months. This consortium (hereafter referred to as ‘the evaluation consortium’) will undertake an independent evaluation of the testing capabilities delivered by the national COVID-19 testing programme in England from October 2020 to March 2022 (hereafter referred to as ‘the evaluation’), with a focus on public health outcomes. This evaluation will seek to capture key learnings from the rollout of testing to the various target populations via the different testing services during this period. The insights gained will inform the formulation of key considerations and recommendations for future pandemic preparedness. Owing to the complexity and rapidly evolving nature of the testing response, no evaluation of the national testing service in England, utilising a mixed methods approach, has been conducted to date.

### Testing services in scope

The evaluation consortium and UKHSA has decided to conduct an overarching evaluation of the national COVID-19 testing programme in England by assessing the combined impact of the asymptomatic and symptomatic testing services. A major component of this will be an evaluation of the universal testing service. This will be complemented by ‘deep-dive’ evaluations of three ‘priority’ testing services (see table 1 for further details):

- Schools (secondary school pupils aged 11 to 18 years)
- Adult social care (staff and residents in care homes)
- Healthcare workers

**Table 1.** Overview of the testing services of the COVID-19 testing programme identified as within scope for this evaluation.

| Testing service | Purpose of testing |
| --- | --- |
| Schools | Testing of staff and students to prevent transmission within schools and, as they are a high-contact group, within the community and to allow schools to continue functioning as normally as possible |
| Adult social care | Testing to prevent outbreaks in vulnerable populations (predominantly those in care homes and those receiving domiciliary care) |
| Healthcare | Testing of NHS staff and patients to reduce the risk of transmission within hospital and other healthcare settings, which could then impact transmission in the community |
| Universal testing | Rapid, asymptomatic, at-home testing with LFDs that were made available to the general public to enable faster detection of cases, thereby allowing more rapid self-isolation |

These four services have been included to ensure a broad spectrum of testing populations in England will be evaluated within the given timeframe, i.e. high volumes (universal testing), high-contact groups (schools) and high-risk groups (care homes and healthcare workers), to best reflect the challenges faced during the pandemic and to balance the findings and recommendations for future pandemics.

The evaluation will follow a hypothesis-led approach, with three key hypotheses developed; based on these hypotheses, the evaluation will seek to answer several research questions (box 1).

##### Box 1. Hypotheses developed and research questions to be explored in the evaluation

**Hypothesis 1:** testing services aimed at protecting high-risk groups, e.g. people in care homes and healthcare workers, led to a reduction in hospitalisations and deaths in these risk groups

**Hypothesis 2:** testing services aimed at high-contact groups, e.g. children in schools, led to a reduction in hospitalisations and deaths in the general population

**Hypothesis 3:** testing services aimed at increasing access to and eligibility for testing and targeting disproportionately impacted groups, e.g. the universal testing service, led to increased testing uptake in these populations

**Research questions:**

1. How was the national COVID-19 testing programme delivered and what factors affected this?
2. What were the barriers and facilitators to access, use and deliver the programme?
3. What were the costs and the cost-effectiveness of the programme?
4. For the universal testing service:
  a. Did the diversity of those reporting test results increase?
  b. Did the barriers and facilitators for testing, reporting and acting on a result change?
5. For each testing service:
  a. Did the service achieve UKHSA’s intended aims and purposes of the service?
  b. Was the service cost-effective?
  c. If testing were to be implemented again, what are the barriers and facilitators to increase access, use and delivery of tests?

## Methodology

### Principles guiding the evaluation

The testing programme can be considered to be a complex public health intervention due to the complexity of the intervention design, which evolved over time according to policy changes and the evolving disease context within which it was implemented.(23) Therefore, this retrospective evaluation will employ a mixed methods approach, utilising existing frameworks that have previously been applied to the evaluation of complex interventions, being broadly divided into process, outcome and impact evaluation components.(24-27)

The evaluation will consist of a phased process:

- A scoping phase, to develop a Theory of Change (ToC), evaluation aims and research questions
- A design phase, to agree the mixed methods approach to the evaluation and confirm process, outcome and impact indicators
- A conduct phase, to collect, review and synthesise data
- A recommend phase, during which findings and data will be triangulated to form conclusions and recommendations

The Medical Research Council (MRC) framework for process evaluation details three key interrelated components required as part of any evaluation of a complex public health process: implementation, mechanisms of impact, and context.(27) The ToC approach(28, 29) – a theory of how and why an initiative works – will therefore be used to map the causal pathways for each of the testing services, as this approach lends itself to understanding complex interventions with multiple causal pathways.(30) The ToC framework will be used to understand the causal pathways and intended and unintended outcomes of each service, as well as exploring the effect of context on each individual testing service’s intended outcomes.

The ToCs will be formulated utilising existing evidence provided by UKHSA and will be subject to further refinement following interviews with UKHSA policy and operational stakeholders. Insights from existing UK government-commissioned evidence in the form of previous COVID-19 service-specific testing evaluations and pilot testing programme data will be utilised where possible to avoid duplication of work. Owing to the unpredictability of evaluating complex interventions such as this, sufficient flexibility will be allowed within the research design to allow emerging research questions to be addressed.(27)

A comprehensive ToC for each of the testing services in scope will subsequently inform process and outcome indicators (table 2) that will be used to assess each service, address the research questions and determine how well each service met its objectives and intended purpose.(31) Based on data availability and granularity, these indicators will be further refined based on what is achievable within the evaluation timeframe.

**Table 2.**
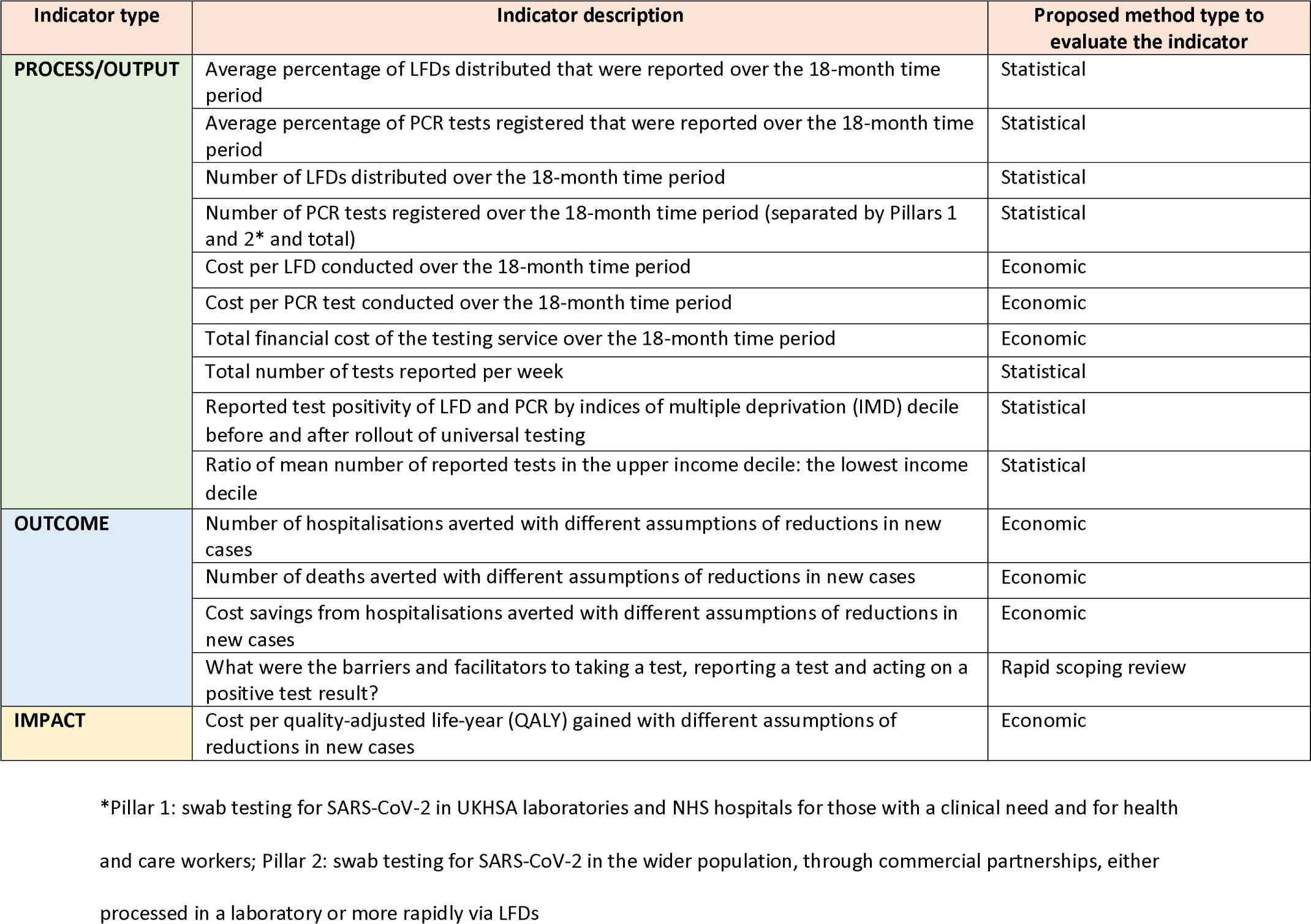
Example of potential evaluation indicators for the overall national testing programme.

The indicators will be categorised as follows:

- Process and output indicators: how did the delivery and uptake of the service compare with what was planned over time, and what factors affected this?
- Outcome indicators: what was the effectiveness of each service in terms of its intended outcomes?
- Impact indicators: what were the broader economic and societal impacts? What were the overall impacts on minimising transmission while limiting harm?

A gap analysis will also be performed during the ‘conduct’ stage to determine whether any crucial data, research or information is unavailable within the existing literature or data sources provided by UKHSA and whether subsequent adjustment of the research questions or descoping is required. This will be decided through a participatory consultation approach with key UKHSA stakeholders.

### Mixed methods approach to be used in the evaluation

A convergent parallel mixed-methods approach will be taken to provide a more holistic understanding of the testing programme, allowing the researchers to capture any unanticipated aspects of testing that may be relevant to the research questions and to explain the quantitative findings.(32)

Findings from the rapid scoping review will be triangulated with the results of the statistical and economic analyses and fed back into the developing ToCs, to refine and explain the indicators, address the hypotheses and help form the recommendations for each testing service during the various evaluation phases. In addition, findings across each of the four testing services will be compared, with the aim of identifying universal as well as service-specific barriers and facilitators to testing, reporting and self-isolation following a positive test result.

The ToCs will be populated in a collaborative manner, enabling the quantitative and qualitative research teams between them to define and align the process and outcome indicators for each testing service. Each team will conduct research concurrently for the various testing services and will communicate their findings at weekly meetings, allowing for emerging findings to be discussed and explored further if deemed necessary. Findings will also be synthesised across the testing services to inform programme-level insights to meet the programme-level research aims and inform the overarching ToC and indicators.

#### Qualitative data analysis: rapid scoping review of testing behaviours

A rapid scoping review will be conducted to evaluate the barriers and facilitators to engaging with COVID-19 testing, reporting results and self-isolating following a positive result in the United Kingdom. This review will aim to 1) provide a summary of the research undertaken on this topic available both in the UKHSA grey literature and in the wider academic literature, 2) identify gaps in research efforts, and 3) provide an overview of key barriers and facilitators for engagement with each testing service, as well for the overall testing programme. A scoping study approach has been selected as the method to synthesise the broad knowledge base on this topic, owing to the large volume of heterogenous literature available and to accommodate the time constraints of the commissioned evaluation.(33) The scoping review will be conducted adhering to the 2005 Arksey and O’Malley framework,(34) incorporating the adaptations proposed in 2010 by Levac and colleagues on iterative study selection and stakeholder consultation (35) and also using the 2015 Joanna Briggs Institute guidance on conducting scoping reviews.(36) The findings will be reported according to the standardised PRISMA-ScR (Preferred Reporting Items for Systematic reviews and Meta-Analyses extension for Scoping Reviews) checklist.(37)

##### Search strategy and the selection of evidence

A wide search strategy has been developed, using key phrases from relevant articles(34) (see table 3 for categories and example terms). This will be used to identify literature that describes behaviour around COVID-19 testing, reporting and self-isolation in the UK during the COVID-19 pandemic. The search strategy will be adapted for each database and information source that is searched and will be refined based on key words in sources identified during the search.

**Table 3.**
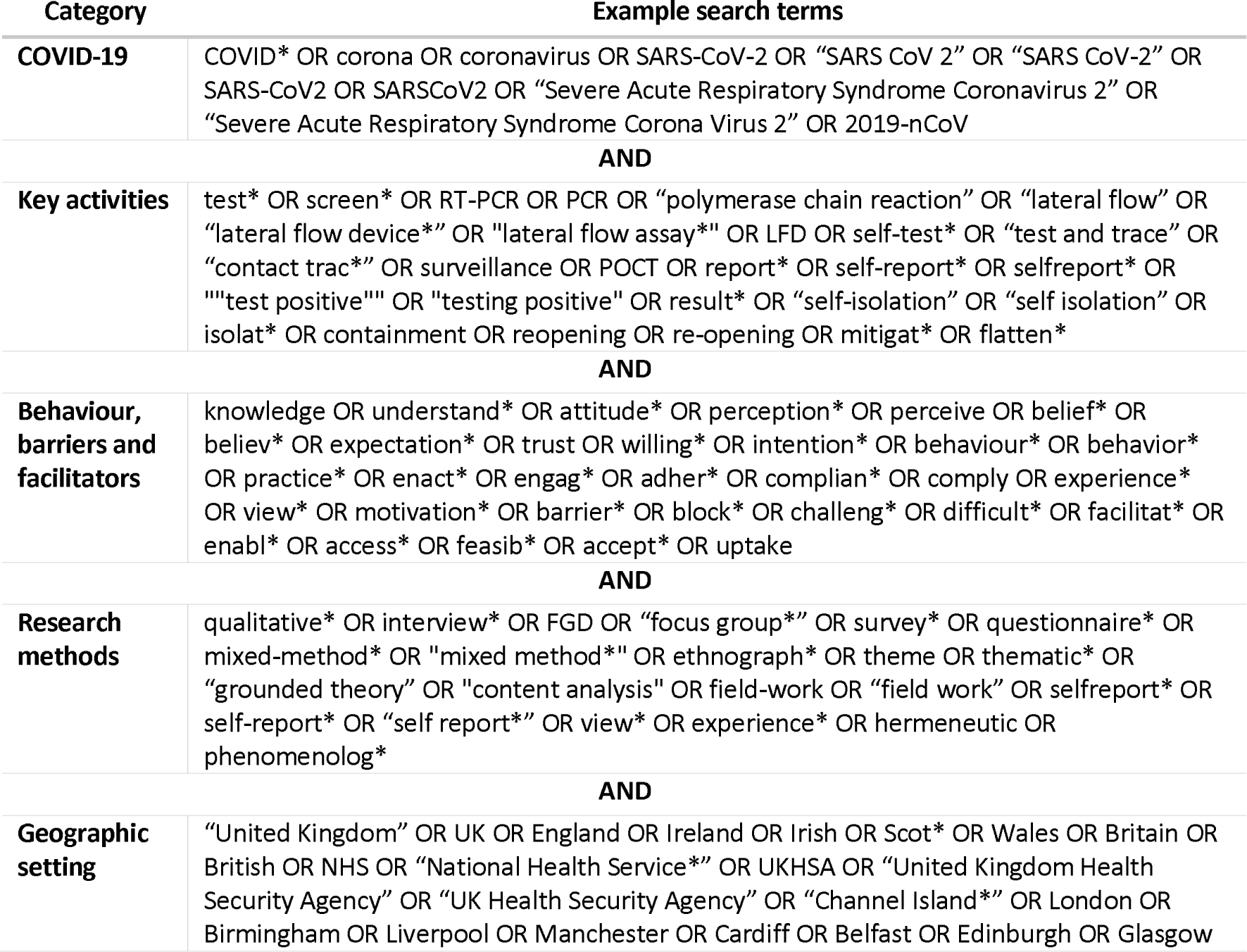
Search categories and examples of search terms.

The databases to be searched include PubMed, Scopus and the WHO COVID-19 Research Database. The search strategy aims to identify both published and unpublished studies (including grey literature), as well as reports and guidance documentation. Research, published from 1 January 2020 onwards and in English, will be included in the review if they focus on one or more of the following three behaviours: undertaking a test, reporting a test result and self-isolating following a positive result, symptoms or a positive contact (see table 4 for search limits and eligibility criteria).

**Table 4.** Summary of the search parameters and limits as well as the inclusion and exclusion criteria, categorised according to the ‘population, context, concept’ search framework. (34, 36)

*Click or tap here to enter text.*
|  | Inclusion | Exclusion |
| --- | --- | --- |
| **SEARCH LIMITS** |  |  |
| **Language** | Published in English | Published in languages other than English |
| **Dates** | Published between 1 January 2020 and the search date (the database search was conducted on 7 November 2022 and the UKHSA documents were received throughout September to December 2022) | Published before 2020 |
| **Methods** | Qualitative or mixed-methods studies; quantitative surveys | Quantitative studies only reporting associations between demographic variables and behavioural outcomes |
| **ELIGIBILITY** |  |  |
| **Literature** | Journal articles, peer-reviewed material, articles under review, published books and book chapters, other academic research, research commissioned by governments, unpublished reports | Opinion or statement pieces, magazine articles, blog posts |
| **Population** | England, Northern Ireland, Scotland, Wales and the islands making up the British Isles; multi-country studies were included if they included one of these settings | Countries outside the UK, including the Republic of Ireland |
| **Concept (key activities)** | Descriptions of individuals’ behaviour and/or barriers to testing and/or facilitators to testing with regards to the following key activities:<br>- Antigen testing for COVID-19 (with a focus on LFDs, but including LAMP and PCR testing)<br>- Reporting test results<br>- Isolating (with a focus on isolating due to a positive COVID-19 test result, but including isolating after being identified as a close contact of a COVID-19-positive case)<br>The description of behaviours included associations of survey responses with behaviour or intention to test, report or isolate | Descriptions of testing, reporting or self-isolation but not the behaviour associated with them, e.g. describes the sensitivity of a specific test<br>Descriptions of testing for antibodies<br>Descriptions of the barriers or facilitators to self-isolation in the context of social distancing, self-isolation if symptomatic or traveller isolation (hotel quarantine)<br>Descriptions of associations of demographic factors with behaviour or intention to test, report or isolate<br>Testing, reporting results or self-isolation after a positive result in the context of other diseases<br>Descriptions of facilitators or |
|  |  | barriers to other COVID-19-related behaviours, such as vaccination or social distancing<br>Descriptions of the impact of testing/reporting/self-isolation on behaviour<br>Knowledge, attitudes or perceptions in relation to COVID-19 itself |

##### Stakeholder-identified supplementary data

UKHSA has identified a repository of data and documentation, comprising more than 5000 reports and internal documents, that may be of potential relevance to the evaluation. Upon commencement of the evaluation, and if a review of the documents highlights further potentially relevant sources, additional documentation will be requested by the evaluation consortium. This documentation will be searched for evidence that supports the consortium’s efforts to understand how the testing services were intended to work, how they were experienced and any prior measurement of their effectiveness. Supplementary documents to be provided by UKHSA could include:

- Testing guidance published by UKHSA
- Testing process documentation
- Business cases
- Primary qualitative or quantitative research (including behavioural studies)
- Documentation involving reporting, managing or measuring the testing programme
- Previous evaluations of testing services

Once all of the publicly available data have been screened, these stakeholder-identified sources will also be reviewed for inclusion.

##### Stakeholder consultation

Stakeholder engagement is considered useful for adding methodological rigour to scoping studies.(35) Therefore, stakeholders from the evaluation sponsor team within UKHSA will be consulted to help identify additional sources of published and unpublished evidence (described above in ‘Supplementary Data’) and offer perspective and validate the findings. Additional sources identified in this way will be included in the scoping review as ‘stakeholder-identified studies’,(38) and insights from these conversations with stakeholders will be anonymised and incorporated into the discussion of the scoping review results. Due to the rapid turnover of testing programme staff members, the availability of stakeholders that the research team will be able to interview is limited. However, the research team has been provided with the names of at least ten policy staff per testing service to interview as part of the stakeholder consultation. This list of names was provided by the funder.

Ethics Committee approval for this research has been granted on the basis that informed consent be sought prior to engaging with any UKHSA stakeholders and that anonymity will be preserved in our write-up of the findings. To preserve anonymity, robust measures will be implemented, including the development of pseudonyms or codes for participants, secure storage of data on UKHSA laptops and careful handling of information during data analysis and reporting. The anonymity preservation strategy has been discussed and refined in collaboration with the study sponsor (UKHSA) and their stakeholders, prior to ethics approval being granted.

##### Data extraction, charting and synthesis

Data extracted from each evidence source will include study metadata (authors, title, year of publication/dissemination, publication stage, country, participant characteristics and methods), the setting (testing service and key activity), and information about the perceptions, experiences and the barriers and facilitators to each of the key activities (testing, reporting and isolating).

All sources will be collated, uploaded into Rayyan, and duplicates removed.(39) Following an initial screening pilot to refine the eligibility criteria, titles and abstracts will then be screened by two reviewers for assessment against the refined inclusion criteria. A sample of ≥20% will be reviewed by a third reviewer to ensure consistency of inclusion.(35) The inter-rater agreement will then be calculated for the final list using Gwet’s first-order agreement coefficient (AC1). (40) Potentially relevant sources will be extracted fully and then assessed against the inclusion criteria. Any disagreements will be resolved through discussion, and with an additional reviewer if no consensus is reached. The same process will be undertaken for the supplementary data.

Once all the data have been extracted and assessed against the inclusion criteria, we will synthesise the data thematically. This process will be repeated for each testing service. Given the rapid timelines, the aim of the work and the scoping study methodology guidance, the included publications will not be assessed for quality.

#### Quantitative data analysis: statistical methods

##### Data collection

Quantitative data will be obtained via the secretariat of the study sponsor within UKHSA, existing UKHSA repositories, the UK Office for National Statistics (ONS), NHS Digital and Public Health Scotland; by applying directly to various holders of non-public datasets; and from other public sources of data where available, such as ISARIC (International Severe Acute Respiratory and Emerging Infection Consortium). General datasets will include SARS-CoV-2 seroprevalence surveys; COVID-19 vaccination data; testing coverage data; and COVID-19 cases, hospitalisations and deaths.

##### Data analysis

The data will be analysed with the aim of 1) providing summaries of indicators identified in the ToCs, in relation to the implementation of each testing service, to better understand the extent and reach of each service and validate findings from the rapid scoping review in relation to behaviours; and 2) providing estimates of the impact of each testing service, which will feed into the cost-effectiveness evaluations. Service-specific statistical approaches will be developed for each of the four testing services.

Specified primary outcome indicators will be defined for each testing service, as defined in the ToCs. Outcomes and appropriate counterfactual comparators will be defined, contingent on obtaining access to relevant data sources; these definitions may be refined using an iterative process based on concurrent analysis of qualitative data from the rapid scoping review. For each testing service, a regression-based approach(24) will be undertaken together with analysis of data at the local authority level (or, if the data allow, at a finer spatial aggregation), accounting for potential confounding factors such as age, sex and ethnicity profiles, as well as indicators of deprivation, population density and relevant chronic illnesses. If determined to be relevant, in collaboration with UKHSA stakeholders, predictors of engagement with health services, such as vaccination uptake or access to internet services, will also be included.

Where available, reported case and prevalence data at the lower tier local authority (LTLA) level will be combined to determine true case-detection ratios, defined as the percentage of all true cases that were captured by the national testing programme, using the statistical debiasing methodology described by Nicholson et al.(41)

#### Quantitative data analysis: economic evaluation methods

##### Data collection

The economic analyses will primarily comprise cost-effectiveness analyses, adopting a provider perspective, incorporating costs to the NHS and to local authorities. For the schools testing service analysis, a societal perspective will be adopted to quantify potential productivity losses. A rapid literature review of publicly available economic data will be conducted using keyword searches of scientific databases, as well as a search of the grey literature using Google Scholar. Data relating to the volumes of tests distributed to the various testing services and the associated costs will be obtained from UKHSA. Data relating to payments made to individuals who were isolating, and other payments made, will be obtained from DHSC. Costs will be apportioned to the four testing services according to the volumes of tests distributed. The costs of hospitalisations will be obtained from the National Schedule of NHS Costs 2020/21.(42)

##### Data analysis

The data will be analysed with the aim of 1) providing an estimate of the costs for each testing service and 2) providing estimates of the value for money of each testing service. The outcome measure of quality-adjusted life-years (QALYs) gained will be used for the economic analyses. The National Institute for Health and Care Excellence (NICE) defines a QALY as ‘a measure of the state of health of a person or group in which the benefits, in terms of length of life, are adjusted to reflect the quality of life. One QALY is equal to 1 year of life in perfect health. QALYs are calculated by estimating the years of life remaining for a patient following a particular treatment or intervention and weighting each year with a quality-of-life score (on a 0 to 1 scale)’.(43) The cost per QALY gained is a critical value that can be used to determine whether an intervention is cost-effective and is used by NICE to determine whether a proposed new treatment can be covered by the NHS. We will use a value of GBP 70,000 as the willingness to pay threshold for interventions for COVID-19, based on HM Treasury’s Green Book (2022).(44) This means that an intervention that costs less than GBP 70,000 per QALY gained can be considered cost-effective. In any graphs produced, we will also indicate the NICE willingness to pay threshold of GBP 30,000, for reference. QALY weights will be obtained from the relevant literature. Sensitivity analyses will be conducted to test the outcomes against the assumptions, using ranges of estimates from the statistical analyses and the literature, for each of the testing services.

### Data management plan

All individuals on the research team will have UK government Baseline Personnel Security Standard clearance.(45) All ‘official-sensitive’ data will be accessed via official, UKHSA-approved secure portals and secure UKHSA laptops. All data to be used in the proposed evaluation will be obtained from UKHSA, allied UK government bodies, such as NHS Digital and the Office for National Statistics (ONS), and independent research organisations.

## Discussion

The national COVID-19 testing programme in England was rapidly set up, scaled, and adapted over time in response to emerging knowledge about COVID-19 transmission, severity and the availability of other public health interventions, such as vaccination. As a result, ongoing monitoring and evaluation of testing as a public health intervention and its effectiveness was not uniformly implemented. This evaluation will therefore aim to retrospectively evaluate the national testing programme in England as a whole, with a deep dive into the schools, adult social care, healthcare worker and universal testing services, using a mixed-methods approach. We believe that the combined strengths of quantitative and qualitative approaches are necessary to evaluate such a complex intervention and will allow for a broader spectrum of insights and triangulation to inform future learnings.

For the purpose of this evaluation, alongside a high-level evaluation of the national testing programme as a whole, we have chosen to narrow our scope to conduct deep dives into four testing services to reflect the spectrum of the impact of testing on both high-risk and high-contact populations and to generate learnings that will be of most value, within the constraints of the evaluation time period.

To the best of the authors’ knowledge, this evaluation will be the first of its kind to produce a portfolio of evidence on testing effectiveness and public health impact for a nationally deployed testing service, focusing on the behavioural, economic, equity and public health impacts. The findings of the evaluation are expected to strengthen the evidence base in relation to COVID-19 testing and identify which aspects when deploying a complex public health intervention, such as testing, will be necessary for decision-makers to prioritise when mitigating future pandemic threats.

## Data Availability

All data produced in the present work are contained in the manuscript

## Ethics statement

This study protocol has been granted ethics approval by the UKHSA Research Ethics and Governance Group (reference NR0347). All relevant ethics guidelines have been followed. Should the research approach or methods change following this version of the protocol, the UKHSA Research Ethics and Governance Group will be notified and the appropriate ethics guidelines followed.

## Disclaimer

This protocol represents research in progress, commissioned and funded by UKHSA, to be performed between August 2022 and March 2023. The scope and depth of testing services covered by this research were pre-agreed with UKHSA and were limited to the availability and provision of data available at the time this protocol was written. The corresponding author has the right to grant on behalf of all authors, and does grant on behalf of all authors, an exclusive licence (or non-exclusive for government employees) on a worldwide basis to the BMJ Publishing Group Ltd to permit this article (if accepted) to be published in BMJ editions and any other BMJ Publishing Group Ltd products and sublicences such use and exploit all subsidiary rights, as set out in the licence. No patients or members of the public were involved in this research.

## Acknowledgements

Scientific writing assistance and editorial support was provided by Adam Bodley, according to Good Publication Practice guidelines. The authors would like to acknowledge the efforts of the UKHSA Scientific Advisory Group, who provided peer review and approval of the study protocol on behalf of the funder.

---

**Appendix A:** EY-Oxford Health Analytics Consortium Membership (in addition to headline authors)

| **Name** | **Affiliation** |
| --- | --- |
| Adam Bodley | Impact Factor Editing, Oxford |
| Ainura Moldokmatova | Big Data Institute, Li Ka Shing Centre for Health Information and Discovery, Nuffield Department of Medicine, University of Oxford |
| Anastasiia Polner | Ernst & Young LLP London |
| Bo Gao | Centre for Tropical Medicine and Global Health, Nuffield Department of Medicine, University of Oxford |
| Caroline Franco | Nuffield Department of Clinical Medicine, University of Oxford \| Institute of Theoretical Physics - São Paulo State University (UNESP) |
| Jared Norman | Modelling and Simulation Hub, Africa (MASHA), University of Cape Town |
| Katie Douglass | Ernst & Young LLP London |
| Liberty Cantrell | Department of Paediatrics, University of Oxford |
| Lok (Joseph) Tsui | Department of Biology, University of Oxford |
| Ma'ayan Amswych | Ernst & Young LLP London |
| Muhammad Kasim | Department of Physics, University of Oxford |
| Randolph Ngwafor Anye | Nuffield Department of Primary Care Health Sciences, University of Oxford |
| Ricardo Aguas | Centre for Tropical Medicine and Global Health, Nuffield Department of Medicine, University of Oxford |
| Richard Creswell | Department of Computer Science, University of Oxford |
| Sabine Dittrich | Centre for Tropical Medicine and Global Health, Nuffield Department of Medicine, University of Oxford |
| Sheetal Silal | Centre for Tropical Medicine and Global Health, Nuffield Department of Medicine, University of Oxford |
| Siyu Chen | Centre for Tropical Medicine and Global Health, Nuffield Department of Medicine, University of Oxford |
| Sompob Saralamba | Centre for Tropical Medicine and Global Health, Nuffield Department of Medicine, University of Oxford |
| Sophie Dickenson | Ernst & Young LLP London |
| Sumali Bajaj | Biodiversity, Ecology & Evolutionary Processes Stream, Department of Biology, University of Oxford |
| Sunil Pokharel | Centre for Tropical Medicine and Global Health, Nuffield Department of Medicine, University of Oxford |
| Tracy Evans | Nuffield Department of Primary Care Health |
|  | Sciences, University of Oxford |
| Umar Mahmood | Ernst & Young LLP London |
| Wirichada Pan-ngum | Centre for Tropical Medicine and Global Health,<br>Nuffield Department of Medicine, University of<br>Oxford |
| Zoe Echanah | Ernst & Young LLP London |

